# UB-612, a Multitope Universal Vaccine Eliciting a Balanced B and T Cell Immunity against SARS-CoV-2 Variants of Concern

**DOI:** 10.1101/2022.04.11.22272364

**Authors:** Chang Yi Wang, Kao-Pin Hwang, Hui-Kai Kuo, Be-Sheng Kuo, Hope Liu, Kuo-Liang Hou, Wan-Yu Tsai, Han-Chen Chiu, Yu-Hsin Ho, Jennifer Cheng, Min-Sheng Wang, Ya-Ting Yang, Po-Yen Chang, Yea-Huei Shen, Wen-Jiun Peng

## Abstract

**Importance:** The SARS-CoV-2 non-spike structural proteins of nucleocapsid (N), membrane (M) and envelope (E) are critical in the host cell interferon response and memory T-cell immunity and have been grossly overlooked in the development of COVID vaccines.

**Objective:** To determine the safety and immunogenicity of UB-612, a multitope vaccine containing S1-RBD-sFc protein and rationally-designed promiscuous peptides representing sequence-conserved Th and CTL epitopes on the Sarbecovirus nucleocapsid (N), membrane (M) and spike (S2) proteins.

**Design, setting and participants:** UB-612 booster vaccination was conducted in Taiwan. A UB-612 booster dose was administered 6-8 months post-2^nd^ dose in 1,478 vaccinees from 3,844 healthy participants (aged 18-85 years) who completed a prior placebo (saline)-controlled, randomized, observer-blind, multi-center Phase-2 primary 2-dose series (100-μg per dose; 28-day apart) of UB-612. The interim safety and immunogenicity were evaluated until 14 days post-booster.

**Exposure:** Vaccination with a booster 3^rd^-dose (100-μg) of UB-612 vaccine.

**Main outcomes and measures:** Solicited local and systemic AEs were recorded for seven days in the e-diaries of study participants, while skin allergic reactions were recorded for fourteen days. The primary immunogenicity endpoints included viral-neutralizing antibodies against live SARS-CoV-2 wild-type (WT, Wuhan strain) and live Delta variant (VNT_50_), and against pseudovirus WT and Omicron variant (pVNT_50_). The secondary immunogenicity endpoints included anti-S1-RBD IgG antibody, S1-RBD:ACE2 binding inhibition, and T-cell responses by ELISpot and Intracellular Staining.

**Results:** No post-booster vaccine-related serious adverse events were recorded. The most common solicited adverse events were injection site pain and fatigue, mostly mild and transient. The UB-612 booster prompted a striking upsurge of neutralizing antibodies against live WT Wuhan strain (VNT_50_, 1,711) associated with unusually high cross-neutralization against Delta variant (VNT_50_, 1,282); and similarly with a strong effect against pseudovirus WT (pVNT_50,_ 6,245) and Omicron variant (pVNT_50_, 1,196). Upon boosting, the lower VNT_50_ and pVNT_50_ titers of the elderly in the primary series were uplifted to the same levels as those of the young adults. The UB-612 also induced robust, durable VoC antigen-specific Th1-oriented (IFN-γ^+^-) responses along with CD8^+^ T-cell (CD107a^+^-Granzyme B^+^) cytotoxicity.

**Conclusions and relevance:** With a pronounced cross-reactive booster effect on B- and T-cell immunity, UB-612 may serve as a universal vaccine booster for comprehensive immunity enhancement against emergent VoCs.

**Trial registration:** [ClinicalTrials.gov: NCT04773067]

**KEY POINTS:** *Question:* Facing ever-emergent SARS-CoV-2 variants and long-haul COVID, can composition-updated new vaccines be constructed capable of inducing striking, durable booster-recalled B/T-immunity to prevent infection by VoCs?

*Findings:* In a Phase-2 extension study, a booster dose of UB-612 multitope protein-peptide vaccine prompted high viral-neutralizing titers against live wild-type virus (VNT_50_, 1,711), Delta variant (VNT_50_, 1,282); pseudovirus wild-type (pVNT_50_, 6,245) and Omicron variant (pVNT_50_, 1,196). Robust, durable Th1-IFNγ^+^ responses and CD8^+^ T cell-(CD107a^+^-Granzyme B^+^) cytotoxic activity were both observed.

*Meaning:* UB-612 RBD-sFc vaccine armed with T cell immunity-promoting conserved N, M and S2 Th/CTL epitope peptides may serve as a universal vaccine to fend off new VoCs.

## INTRODUCTION

The devastation of the virulent Delta and the hyper-transmissible, pathologically-lesser Omicron variants (**eFigure 1 in the Supplement**) has declined and COVID restrictions have been lifted. While mRNA booster vaccination could reverse Omicron-induced decrease of serum neutralizing antibodies^1-11^ and reduce rates of hospitalization and severe disease, it offers less protection against infection and mild disease. Still, Omicron can cause breakthrough infections even after the fourth vaccine jab.^11^

Omicron-infected asymptomatic and mild cases shall not be treated akin to a flu. Reportedly, COVID can take a serious toll on heart health,^12^ presumably to stay as a part of long-haul COVID.^13^ This toll, beyond the myocarditis and pericarditis associated with mRNA vaccines,^14^ encompasses a wide range of inflammatory cardiovascular disorders that elevates, depending on COVID severity, from asymptomatic, symptomatic, to acute infection cases.^12,15,16^

Facing ever-emergent variants and long-haul COVID, new composition-updated vaccines that can prevent infection are strongly advocated.^17,18^ A universal vaccine capable of inducing durable cross-reactive viral-neutralizing antibodies along with broad T-cell immunity is desirable. Apparently, the currently authorized spike-only vaccines do not incorporate SARS-CoV-2’s non-spike structure proteins of envelope (E), membrane (M) and nucleocapsid (N), the regions critically involved in the host cell interferon response and T-cell memory^19-22^ yet grossly overlooked in COVID vaccine development.

Here we report the booster immunogenicity and safety in Phase-2 trial of UB-612, a multitope vaccine^23^ (**Figure 1A**) containing an S1-RBD-sFc protein and rationally-designed Th and CTL epitope peptides on the Sarbecovirus nucleocapsid (N), membrane (M) and spike (S2) proteins that are sequence-conserved across all VoCs (**Figure 1B**).

**Figure 1.**
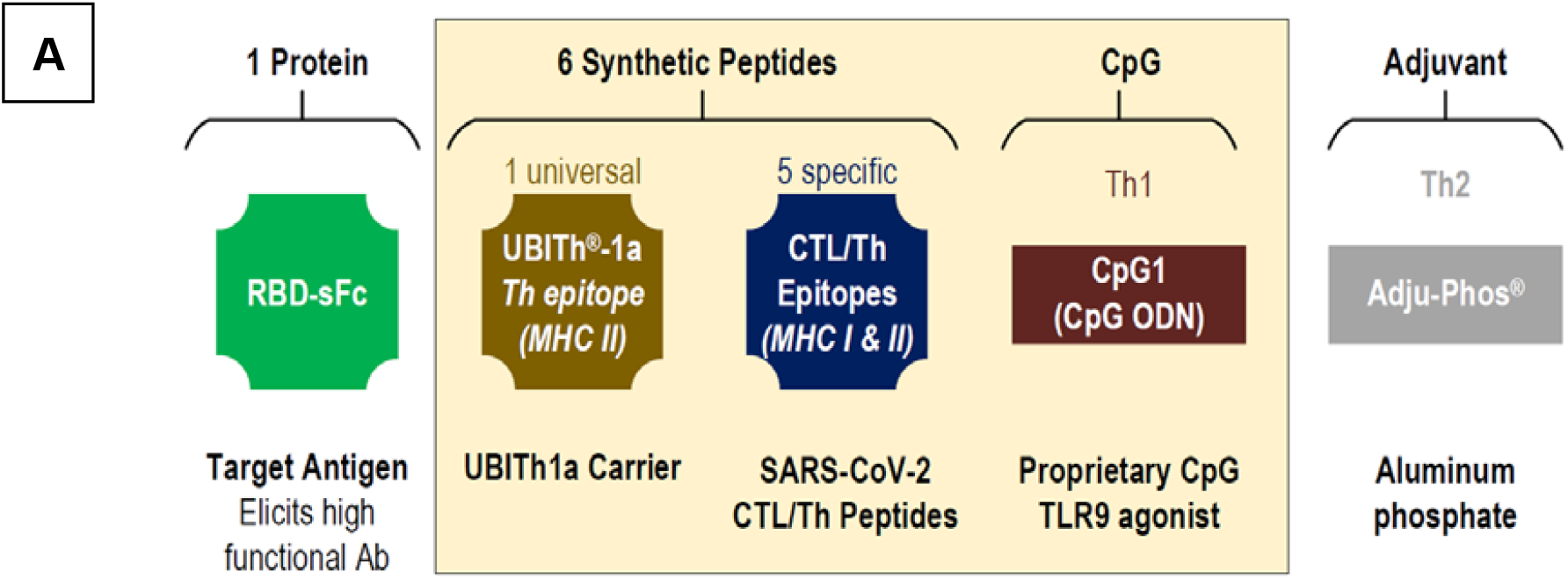
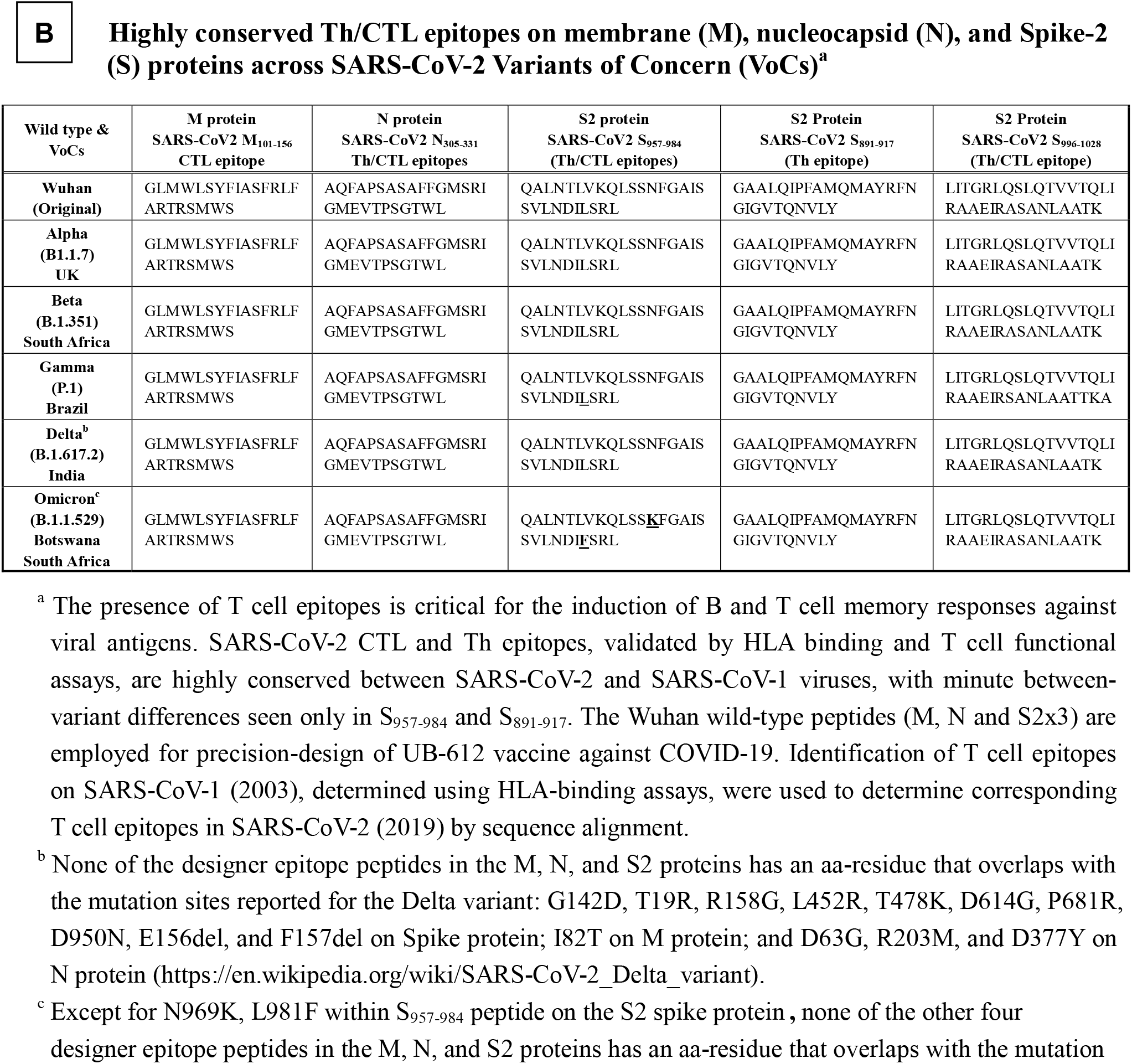
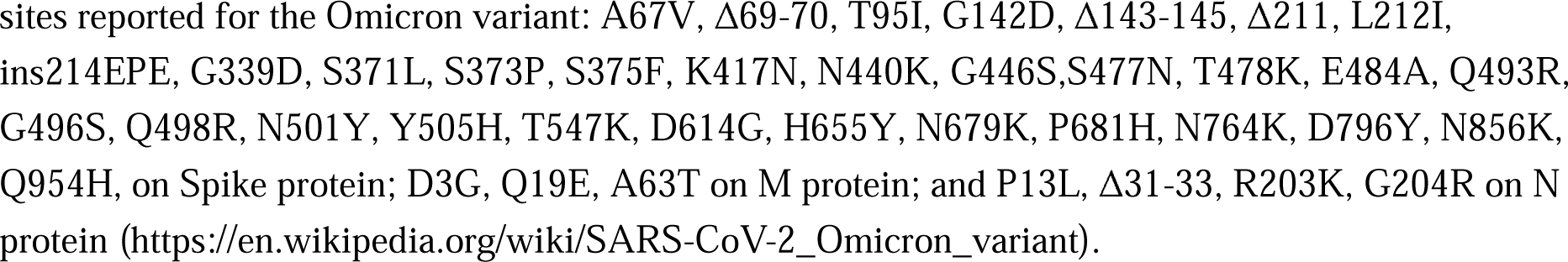
Components of UB-612 multitope protein-peptide vaccine with highly conserved T cell epitope peptides to achieve balanced humoral and cellular immunity. UB-612 vaccine construct contains an S1-RBD-sFc single chain fusion protein targeting only the receptor ACE2 conformational binding site for the B cell epitopes, plus five synthetic Th/CTL peptides derived from SARS-CoV-2 nucleocapsid (N), membrane (M) and spike S2 proteins, and the proprietary UBITh1a peptide as a T cell activation catalyst, all with promiscuous MHC (HLA) class I and II binding motifs with broad MHC (HLA) genetic coverage, and known to bind and trigger T cell proliferation. These positively charged designer T peptides are bound to our proprietary highly negatively charged CpG1 through charge neutralization, which is then bound to Adjuphos^®^ as an adjuvant to constitute the UB-612 vaccine drug product.

## METHODS

### Design of Extension Booster Trial, Objectives, and Oversight

#### Booster 3rd-dose following the Phase-2 trial primary 2-dose series

We conducted a booster vaccination study (n = 1,478) which was an extension arm of the phase-2, placebo-controlled, randomized, observer-blind, multi-center primary 2-dose study (**eFigure 2A in the Supplement**) [ClinicalTrials.gov: NCT04773067] in Taiwan with 3,844 healthy male or female adults aged >18 to 85 years (**eFigure 2B in the Supplement**) who received two intramuscular doses (28 days apart) of 100 μg UB-612 or saline placebo.^24^ The objectives of the third-dose extension study was to determine the booster-induced safety and immunogenicity after unblinding, 6 to 8 months after the second dose.

The Principal Investigators at the study sites agreed to conduct the study according to the specifics of the study protocol and the principles of Good Clinical Practice (GCP); and all the investigators assured accuracy and completeness of the data and analyses presented. The protocols were approved by the ethics committee at the site and all participants provided written informed consent. Full details of the booster trial design, inclusion and exclusion criteria, conduct, oversight, and statistical analysis plan are available in **Supplement**.

### Vaccine Product and Placebo

Like the same vaccine product used in the Phase-1 primary and booster and the Phase-2 primary trials, UB-612 vaccine used in the present phase-2 extension booster vaccination is a multitope vaccine designed to activate both humoral and cellular responses. For SARS-CoV-2 immunogens, UB-612 combines a CHO-expressed S1-RBD-sFc fusion protein (Wuhan strain) and a mixture of synthetic T helper (Th) and cytotoxic T lymphocyte (CTL) epitope peptides, which are selected from immunodominant M, S2 and N regions known to bind to human major histocompatibility complexes (MHC) I and II. The preparation of UB-612 vaccine product consists of compounding, filtration, mixing, and filling operations. Before addition of the subunit protein S1-RBD-sFc, the individual components of the vaccine are filtered through a 0.22 micron membrane filter, including the peptide solution (2 µg/mL), CpG1, a proprietary oligonucleotide (ODN), solution (2 µg/mL), 10X protein buffer containing 40 mM Histidine, 500 mM Arginine and 0.6% Tween 80, 20% sodium chloride stock solution. After sequentially addition of each component, the S1-RBD-sFc fusion protein and peptides are formulated with components described as above to form a protein-peptide complex and then is adsorbed to aluminum phosphate (Adju-Phos®) adjuvant. The last step would be addition of water for injection containing the 2-phenoxyethanol preservative solution to make final drug product at 200 µg/mL. The UB-612 vaccine product is stored at 2 to 8 °C. Placebo used in the Phase-2 trial was sterile 0.9% normal saline.

### Trial Procedures and Safety

#### Phase-2 trial of primary and booster third-dose series

The primary safety endpoints the Phase-2 trial were to evaluate the safety and tolerability of all participants receiving study intervention from Days 1 to 365. Vital signs were assessed before and after each injection. Participants were observed for 30 minutes after each injection for changes in vital signs or any acute anaphylactic reactions. After each injection, participants had to record solicited local and systemic AEs in their self-evaluation e-diary for up to seven days while skin allergic reactions were recorded in their e-diary for up to fourteen days. Safety endpoints include unsolicited AEs reported for Days 1 to Day 57 in this interim Phase-2 report. Complete details for solicited reactions are provided in the study protocols.

### Scope of Immunogenicity Investigation

The primary immunogenicity endpoints were the geometric mean titers (GMT) of neutralizing antibodies against SARS-CoV-2 wild-type (WT, Wuhan strain), and the post-booster effects against Omicron and Delta variant were explored as well. For WT and Delta strains, viral-neutralizing antibody titers that neutralize 50% (VNT50) of live SARS-CoV-2 WT and Delta variant were measured by a cytopathic effect (CPE)-based assay using Vero-E6 (ATCC® CRL-1586) cells challenged with SARS-CoV-2-Taiwan-CDC#4 (Wuhan strain) and SARS-CoV-2-Taiwan-CDC#1144 (B.1.617.2; Delta variant). The replicating virus neutralization test conducted at Academia Sinica was fully validated using internal reference controls and results expressed as VNT_50_. The WHO reference standard was also employed and results reported in international units (IU/mL). For WT and Omicron strains, 50% pseudovirus neutralization titers (pVNT_50_) were measured by neutralization assay using HEK-293T-ACE2 cells challenged with SARS-CoV-2 pseudovirus variants expressed the spike protein of WT and Omicron BA.1variants. Assay methods are detailed below.

The secondary immunogenicity endpoints include anti-S1-RBD IgG antibody, inhibitory titers against S1-RBD:ACE2 interaction, and T-cell responses assayed by ELISpot and Intracellular Staining. The RBD IgG ELISA was fully validated using internal reference controls and results expressed in end-point titers. The WHO reference standard was also employed and results reported in Binding Antibody Units (BAU/mL) (70, Lancet 2021;397(10282):1347-1348.). A panel of 20 human convalescent serum samples from COVID-19 Taiwan hospitalized patients aged 20 to 55 years were also tested for comparison with those in the vaccinees. Human peripheral blood mononuclear cells (PBMCs) were used for monitoring T cell responses. All bioassay methods are detailed below.

### Viral-neutralizing antibody titers against SARS-CoV-2 wild-type and variants by CPE based live virus neutralization assay

Neutralizing antibody titers were measured by CPE-based live virus neutralization assay using Vero-E6 cells challenged with wild type (SARS-CoV-2-Taiwan-CDC#4, Wuhan) and Delta variant (SARS-CoV-2-Taiwan-CDC#1144, B.1.617.2), which was conducted in a BSL-3 lab at Academia Sinica, Taiwan. Vero-E6 (ATCC® CRL-1586) cells were cultured in DMEM (Hyclone) supplemented with 10% fetal bovine serum (FBS, Gibco) and 1x Penicillin-Streptomycin solution (Thermo) in a humidified atmosphere with 5% CO_2_ at 37°C. The 96-well microtiter plates are seeded with 1.2×10^4^ cells/100 μL/well. Plates are incubated at 37° C in a CO_2_ incubator overnight. The next day tested sera were heated at 56 °C for 30 min to inactivate complement, and then diluted in DMEM (supplemented with 2% FBS and 1x Penicillin/Streptomycin). Serial 2-fold dilutions of sera were carried out for the dilutions. Fifty μL of diluted sera were mixed with an equal volume of virus (100 TCID50) and incubated at 37°C for 1 hr. After removing the overnight culture medium, 100 μL of the sera-virus mixtures were inoculated onto a confluent monolayer of Vero-E6 cells in 96-well plates in triplicate. After incubation for 4 days at 37 °C with 5% CO_2_, the cells were fixed with 10% formaldehyde and stained with 0.5% crystal violet staining solution at room temperature for 20 min. Individual wells were scored for CPE as having a binary outcome of ‘infection” or ‘no infection’. Determination of SARS-CoV-2 virus specific neutralization titer was to measure the neutralizing antibody titer against SARS-CoV-2 virus based on the principle of VNT50 titer (≥50% reduction of virus-induced cytopathic effects). Virus neutralization titer of a serum was defined as the reciprocal of the highest serum dilution at which 50% reduction in cytopathic effects are observed and results are calculated by the method of Reed and Muench.

### Neutralizing Antibody Titers against Omicron strain by pesudovirus luciferase assay

Neutralizing antibody titers were measured by neutralization assay using HEK-293T-ACE2 cells challenged with SARS-CoV-2 pseudovirus variants. The study was conducted in a BSL2 lab at RNAi core, Biomedical Translation Research Center (BioTReC), Academia Sinica. Human embryonic kidney (HEK-293T/17; ATCC® CRL-11268™) cells were obtained from the American Type Culture Collection (ATCC). Cells were cultured in DMEM (Gibco) supplemented with 10% fetal bovine serum (Hyclone) and 100 U/mL of Penicillin-Streptomycin solution (Gibco), and then incubated in a humidified atmosphere with 5% CO2 at 37 °C. HEK-293T-ACE2 cells were generated by transduction of VSV-G pseudotyped lentivirus carrying human ACE2 gene. To produce SARS-CoV-2 pseudoviruses, a plasmid expressing C-terminal truncated wild-type Wuhan-Hu-1 strain SARS-CoV-2 spike protein (pcDNA3.1-nCoV-SΔ18) was co-transfected into HEK-293T/17 cells with packaging and reporter plasmids (pCMVΔ8.91, and pLAS2w.FLuc.Ppuro, respectively) (BioTReC, Academia Sinica), using TransIT-LT1 transfection reagent (Mirus Bio). Site-directed mutagenesis was used to generate the Omicron BA.1variants by changing nucleotides from Wuhan-Hu-1 reference strain. For BA.1 variant, the mutations of spike protein are A67V, Δ69-70, T95I, G142D/Δ143-145, Δ211/L212I, ins214EPE, G339D, S371L, S373P, S375F, K417N, N440K, G446S, S477N, T478K, E484A, Q493R, G496S, Q498R, N501Y, Y505H, T547K, D614G, H655Y, N679K, P681H, N764K, D796Y, N856K, Q954H, N969K, L981F. Indicated plasmids were delivered into HEK-293T/17 cells by using TransITR-LT1 transfection reagent (Mirus Bio) to produce different SARS-CoV-2 pseudoviruses. At 72 hours post-transfection, cell debris were removed by centrifugation at 4,000 xg for 10 minutes, and supernatants were collected, filtered (0.45 μm, Pall Corporation) and frozen at −80 °C until use. HEK-293-hACE2 cells (1×10^4^ cells/well) were seeded in 96-well white isoplates and incubated for overnight. Tested sera were heated at 56°C for 30 min to inactivate complement, and diluted in medium (DMEM supplemented with 1% FBS and 100 U/ml Penicillin/Streptomycin), and then 2-fold serial dilutions were carried out for a total of 8 dilutions. The 25 μL diluted sera were mixed with an equal volume of pseudovirus (1,000 TU) and incubated at 37 °C for 1 hr before adding to the plates with cells. After 1-hr incubation, the 50 μL mixture added to the plate with cells containing with 50 μL of DMEM culture medium per well at the indicated dilution factors. On the following 16 hours incubation, the culture medium was replaced with 50 μL of fresh medium (DMEM supplemented with 10% FBS and 100 U/ml Penicillin/Streptomycin). Cells were lysed at 72 hours post-infection and relative light units (RLU) was measured by using Bright-GloTM Luciferase Assay System (Promega). The luciferase activity was detected by Tecan i-control (Infinite 500). The percentage of inhibition was calculated as the ratio of RLU reduction in the presence of diluted serum to the RLU value of virus only control and the calculation formula was shown below: (RLU ^Control^ - RLU ^Serum^) / RLU ^Control^. The 50% protective titer (NT50 titer) was determined by Reed and Muench method.

### Inhibition of RBDWT binding to ACE2 by ELISA

The 96-well ELISA plates were coated with 2 µg/mL ACE2-ECD-Fc antigen (100 μL/well in coating buffer, 0.1M sodium carbonate, pH 9.6) and incubated overnight (16 to 18 hr) at 4 °C. Plates were washed 6 times with Wash Buffer (25-fold solution of phosphate buffered saline, pH 7.0-7.4 with 0.05% Tween 20, 250 μL/well/wash) using an Automatic Microplate Washer. Extra binding sites were blocked by 200 μL/well of blocking solution (5 N HCl, Sucrose, Triton X-100, Casein, and Trizma Base). Five-fold dilutions of immune serum or a positive control (diluted in a buffered salt solution containing carrier proteins and preservatives) were mixed with a 1:100 dilution of RBDWT-HRP conjugate (horseradish peroxidase-conjugated recombinant protein S1-RBD-His), incubated for 30±2 min at 25±2 °C, washed and TMB substrate (3,3’,5,5’-tetramethylbenzidine diluted in citrate buffer containing hydrogen peroxide) is added. Reaction is stopped by stop solution (diluted sulfuric acid, H_2_SO_4,_ solution, 1.0 M) and the absorbance of each well is read at 450nm within 10 min using the Microplate reader (VersaMax). Calibration standards for quantitation ranged from 0.16 to 2.5 μg/mL. Samples with titer value below 0.16 μg/mL were defined as being half of the detection limit. Samples with titer exceed 2.5 μg/mL were further diluted for reanalysis.

### Anti-S1-RBD_WT_ binding IgG antibody by ELISA

The 96-well ELISA plates were coated with 2 µg/mL recombinant S1-RBD_WT_-His protein antigen (100 µL/well in coating buffer, 0.1 M sodium carbonate, pH 9.6) and incubated overnight (16 to 18 hr) at room temperature. One hundred μL/well of serially diluted serum samples (diluted from 1:20, 1:1,000, 1:10,000 and 1:100,000, total 4 dilutions) in 2 replicates were added and plates are incubated at 37 °C for 1 hr. The plates were washed six times with 250 μL Wash Buffer (PBS-0.05% Tween 20, pH 7.4). Bound antibodies were detected with HRP-rProtein A/G at 37 °C for 30 min, followed by six washes. Finally, 100 μL/well of TMB (3,3’,5,5’-tetramethylbenzidine) prepared in Substrate Working Solution (citrate buffer containing hydrogen peroxide) was added and incubated at 37 °C for 15 min in the dark, and the reaction stopped by adding 100 μL/well of H_2_SO_4,_ 1.0 M. Sample color developed was measured on ELISA plate reader (Molecular Device, VersaMax). UBI® EIA Titer Calculation Program was used to calculate the relative titer. The anti-S1-RBD antibody level is expressed as Log_10_ of an end point dilution for a test sample (SoftMax Pro 6.5, Quadratic fitting curve, Cut-off value 0.248).

### T cell responses by ELISPOT

Human peripheral blood mononuclear cells (PBMCs) were used in the detection of the T cell response. For the booster-series third-dose series extension study, ELISpot assays were performed using the human IFN-γ/IL-4 FluoroSpot^PLUS^ kit (MABTECH). Aliquots of 250,000 PBMCs were plated into each well and stimulated, respectively, with 10 μg/mL (each stimulator) of RBD-_WT_+Th/CTL, Th/CTL, or Th/CTL pool without UBITh1a (CoV2 peptides), and cultured in culture medium alone as negative controls for each plate for 24 hours at 37 °C with 5% CO2. The analysis was conducted according to the manufacturer’s instructions. Spot-forming units (SFU) per million cells was calculated by subtracting the negative control wells.

### Intracellular Cytokine Staining (ICS)

Intracellular cytokine staining and flow cytometry was used to evaluate CD4^+^ and CD8^+^ T cell responses. PBMCs were stimulated, respectively, with S1-RBD-His recombinant protein plus with Th/CTL peptide pool, Th/CTL peptide pool only, CoV2 peptides, PMA + Inonmycin (as positive controls), or cultured in culture medium alone as negative controls for 6 hours at 37°C with 5% CO_2_. Following stimulation, cells were washed and stained with viability dye for 20 minutes at room temperature, followed by surface stain for 20 minutes at room temperature, cell fixation and permeabilization with the BD cytofix/cytoperm kit (Catalog # 554714) for 20 minutes at room temperature, and then intracellular stain for 20 minutes at room temperature. Intracellular cytokine staining of IFN-γ, IL-2 and IL-4 was used to evaluate CD4^+^ T cell response. Intracellular cytokine staining of IFN-γ, IL-2, CD107a and Granzyme B was used to evaluate CD8^+^ T cell responses. Upon completion of staining, cells were analyzed in a FACSCanto II flow cytometry (BD Biosciences) using BD FACSDiva software.

### Statistics

For the Phase-2 extension booster vaccination study, the immunogenicity results for Geometric Mean Titer (GMT) are presented with the 95% confidence intervals. Statistical analyses were performed using SAS® Version 9.4 (SAS Institute, Cary, NC, USA) or Wilcoxon sign rank test. Spearman correlation was used to evaluate the monotonic relationship between non-normally distributed data sets. For the Phase-2 primary 2-dose series, the sample size of the trial design meets the minimum safety requirement of 3000 study participants in the vaccine group with healthy adults, as recommended by the US FDA and WHO.

US Food and Drug Administration, Emergency use authorization for vaccines to prevent COVID-19: Guidance for industry, https://downloads.regulations.gov/FDA-2020-D-1137-0019/attachment_1.pdf;

WHO Guidelines on clinical evaluation of vaccines: regulatory expectations, https://cdn.who.int/media/docs/default-source/prequal/vaccines/who-trs-1004-web-annex-9.pdf?sfvrsn=9c8f4704_2&download=true.

## RESULTS

### Phase-2 Booster Trial Population, Reactogenicity and Safety

#### Trial population

After unblinding of Phase-2 trial, 1,478 of the 3,844 healthy study participants who completed the 2-dose primary series (28 days apart) of 100-μg UB-612 (**eFigure 2A in the Supplement**) were enrolled to receive an additional 100-μg booster 3rd-dose 6 to 8 months after the second shot. The booster vaccinees were followed for 14 days for assessment of safety and immunogenicity. The vast majority of participants were of Taiwanese origin, with two groups aged 18-65 years (76%) and 65-85 years (24%) (**eFigure 2B in the Supplement**).

#### Reactogenicity and safety

No vaccine-related serious adverse events (SAEs) were recorded; the most common solicited AEs were injection site pain and fatigue, mostly mild and transient (**eFigure 3 in the Supplement**). The incidence of solicited local AEs slightly increased post-booster (**eFigure 3A in the Supplement**), mostly pain at the injection site (mild, 54%; moderate, 7%). The incidence of skin allergic reaction at post-booster was similar to post-dose 2 (**eFigure 3B in the Supplement**). Fatigue/tiredness, muscle pain, and headaches that belonged to solicited systemic AEs were mostly mild (**eFigure 3C in the Supplement**). Overall, no safety concerns were identified with UB-612.

### Durable T Cells Responses Induced by Th/CTL Epitope Peptides

#### ELISpot

Vaccinees’ peripheral blood mononuclear cells (PBMCs) were collected for evaluation of Interferon-γ^+^ (IFN-γ^+^)-ELISpot. On Day 57 (28 days post-2^nd^ dose), IFN-γ SFU (Spot Forming Unit)/10^6^ cells under stimulation with RBD+Th/CTL increased from the baseline 1.0 to a high 374 SFU (**Figure 2A**), which maintained strong at 261 (70%) at pre-boosting (6-8 months post-2^nd^ dose), then rose to 444 SFU 14 days post-booster (**Figure 2D**). Similar IFN-γ profiles were observed for that stimulated with Th/CTL alone, which increased from the baseline 1.3 to a high 322 SFU on Day 57 (**Figure 2A**), maintained at 182 SFU (∼57%) at pre-boosting and remained strong at 317 SFU 14 days post-booster (**Figure 2D**). Apparently, T cell responses persisted robust (60-70% of the high peak at Day 57) long over 6-8 months.

**Figure 2.**
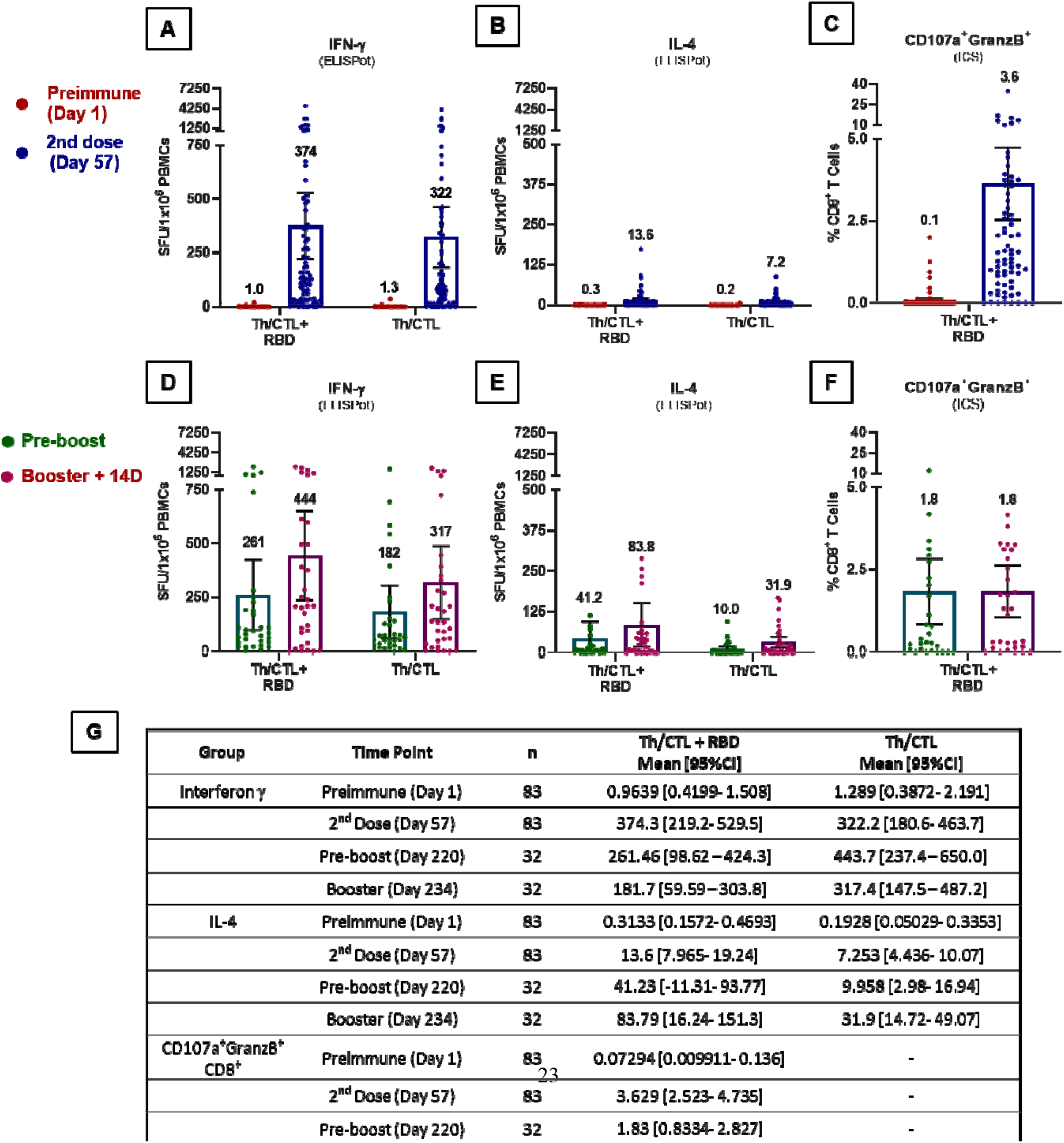
UB-612 induced T cell responses measured by ELISpot and ICS analyses. T-cell responses to stimulation by epitope peptides (Th/CTL+RBD or Th/CTL alone) were analysed with PBMCs collected from 83 vaccinees from Immunogenicity group (n = 83) on Days 57 (28 days after 2^nd^ booster); and from 32 vaccinees from the Immunogenicity (n = 18) or Safety groups (n = 14) who joined the Phase-2 extension booster study to evaluate the T-cell responses in PBMCs on Days 197 to 242 (pre-boosting days) and Days 211 to 256 (14 days post-booster third dose). T-cell responses were measured by **(A, D)** IFN-γ and **(B, E)** IL-4 ELISpot at 10-ug/mL per stimulator. Spot-forming units (SFU) per 1×10^6^ PBMCs producing IFN-γ and IL-4 after stimulation with the RBD+Th/CTL peptide pool or the Th/CTL peptide pool are expressed. The PBMC samples stimulated with Th/CTL+RBD were also evaluated for T cell responses by Intracellular Staining (ICS) (**C, F**), by which the frequency of CD8^+^ T cells producing the measured cytokines (CD107a and Granzyme B) in response to the stimulation of RBD+Th/CTL peptide poolare shown. (**G**) Summary of mean and 95% CI are presented for plots as shown in Figures **(A)** to **(F)**. Horizontal bars indicate mean with 95% CI.

These results indicate that UB-612 can induce a strong, durable IFN-γ^+^ T cell immunity in the primary series and prompt a high level of memory recall upon boosting, and that the presence of Th/CTL peptides is essential and principally responsible for the bulk of the T cell responses, while S1-RBD plays only a minor role. Together with the insignificant low levels of the IL-4^+^ ELISpot responses (**Figures 2B and 2E**), UB-612 vaccination at both primary series and homologous-boosting could induce pronounced Th1-predominant immunity.

#### Intracellular Staining (ICS)

Along with high levels of ELISpot-based T cell responses, CD8^+^ T cells expressing cytotoxic markers, CD107a and Granzyme B, were observed in the primary series, accounting for a remarkable 3.6% of circulating CD8^+^ T cells after re-stimulation with S1-RBD+Th/CTL (**Figure 2C**), which persisted at a substantial 1.8% upon booster vaccination (**Figure 2F**). Apparently, CD8^+^ T cell responses persisted robustly (50% of the high peak at Day 57) long over 6-8 months as well. This suggests a potential of robust cytotoxic CD8^+^ T responses in favor of viral clearance once infection occurs.

### Cross-reactive Neutralizing Antibodies against SARS-CoV-2 Omicron and Delta Variants

#### Anti-Omicron assay with pseudovirus

Based on pseudovirus neutralization assay and observed across all groups aged 18-85 years (n = 41), the UB-612 booster elicited a high geometric mean 50% neutralizing titer (pVNT_50_) at 6,245 against wild-type Wuhan strain (WT) versus Omicron variant at 1,196, representing a 5.2-fold reduction (**Figure 3A**). There was no significant age-dependent booster-induced neutralization effect between young adults (18-65 years) and the elderly (65-85 years) with respect to either anti-WT or anti-Omicron pVNT_50_ level (**Figure 3B**). Both age groups showed a 5.0- to 6.0-fold reduction for anti-Omicron relative to anti-WT, yet the pVNT_50_ of 1,196 still represents a potent viral-neutralizing strength.

**Figure 3.**
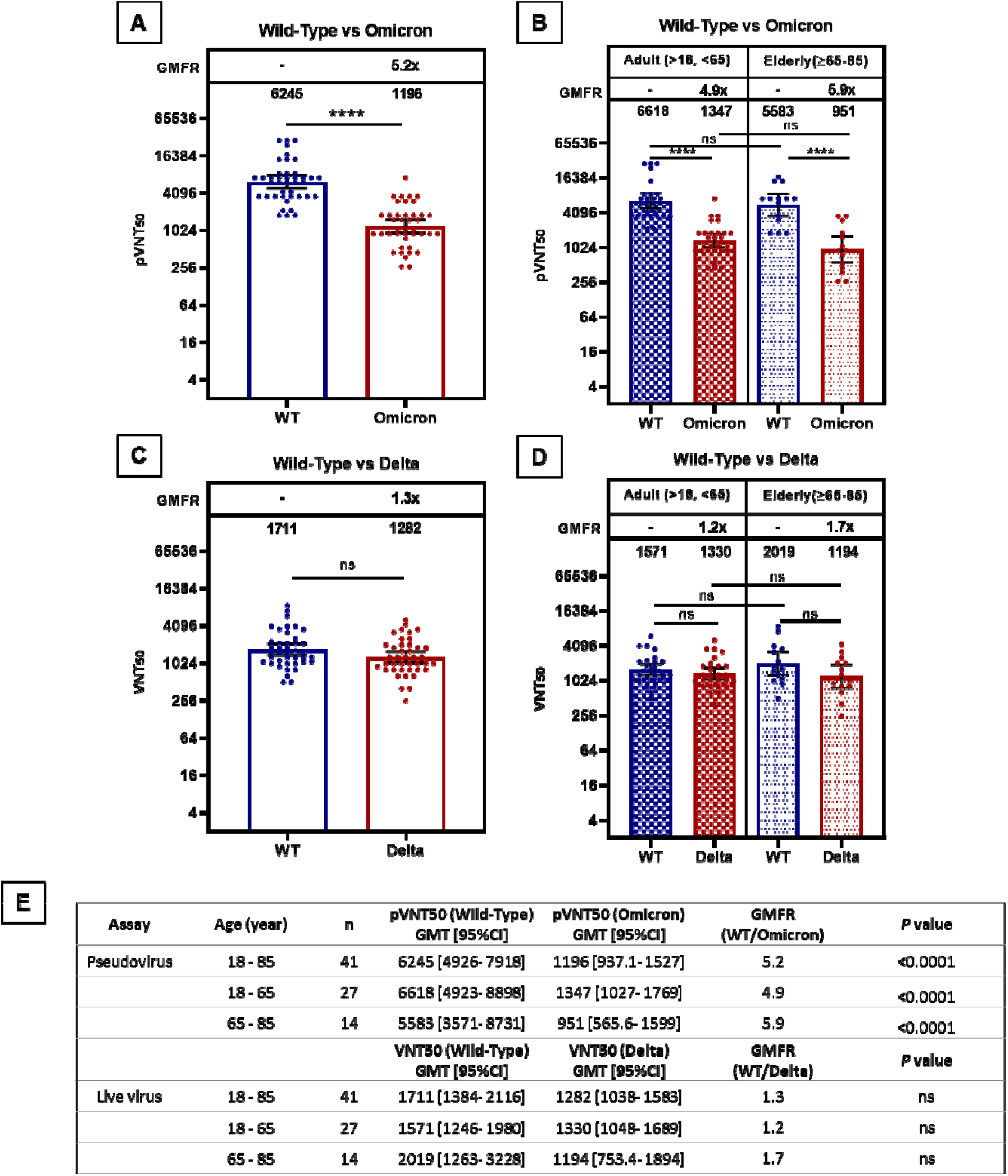
UB-612 booster induced potent neutralization effects against both Omicron and Delta variants. A total of 41 serum samples (n = 27 for 18-65 years; n = 14 for 65-85 years) from 14 days post-booster were subjected to a pseudovirus-luciferase neutralization assay for Omicron and a live virus CPE neutralization assay for Delta variant. The viral-neutralizing antibody geometric mean titers (GMT, 95% CI) that inhibit 50% of SARS-CoV-2 wild-type Wuhan strain (WT), Omicron and Delta were measured. Statistical analysis was performed by the Student’s t-test (ns, *p*>0.05; ****, *p*<0.0001). **(A)** The pVNT_50_ titers 14 days post-booster reached at 6245 against SARS-CoV-2 WT, and at 1196 against Omicron. The 5.2-fold reduction stands for 19% preservation of neutralization titers relative the Wuhan wild type. **(B)** The pVNT_50_ titers are presented by two age groups. **(C)** The VNT_50_ titers 14 days post-booster reached at 1711 against live WT, and at 1282 against Delta. The 1.3-fold reduction stands for 75% preservation of neutralization titer relative to the WT. **(D)** The VNT_50_ titers are presented by two age groups. **(E)** Summary of geometric mean titers (GMT) and 95% CI for plots shown in Figures **(A)** to **(D)**.

#### Anti-Delta assay with live virus

Based on live virus neutralization assay observed across all groups aged 18-85 years (n = 41), the UB-612 booster elicited a geometric mean 50% neutralizing titer (VNT_50_) against wild-type Wuhan strain (WT) at 1,711 versus Delta variant at 1,282, representing a 1.3-fold reduction (**Figure 3C**). Again, there was no significant age-dependent booster-induced neutralization effect between young adults (18-65 yrs.) and the elderly (65-85 yrs.) observed with respect to either anti-WT or anti-Delta VNT_50_ level (**Figure 3D**). Both age groups showed a 1.2- to 1.7-fold reduction for anti-Delta relative to anti-WT.

### Functional correlations between ACE2:RBD_WT_ binding inhibition and viral neutralization

#### Immunogenicity overview, antigenic and functional

Of the 871 Phase-2 study participants enrolled for the primary 2-dose series grouped for Immunogenicity investigation, 302 participants had their sera collected at pre-boosting and 14 days post-booster for antigenic assay by anti-S1-RBD IgG ELISA, and functional assays by ACE2:RBD_**WT**_ binding inhibition ELISA and by neutralization against live SARS-CoV-2 wild-type Wuhan strain by cell-based CPE method (**eFigure 4 in the Supplement**). The results showed pronounced booster-enhanced increases in both target binding and inhibition/neutralization titers by respective 16- to 45-folds.

#### Potent and durable viral-neutralization and ACE2:RBD_WT_ binding inhibition

In the Phase-2 extension booster Immunogenicity group, the immune sera from 87 participants available on Day 1 (pre-dose), Day 57 (28 days post-2^nd^ dose), Day 220 (pre-boosting, 6 to 8 months post-2^nd^ dose), and Day 234 (14 days post-booster) were assayed for neutralization against live WT strain, which showed a high post-booster VNT_50_ titer of 738 (**Figure 4A**), a 17- fold increase over the pre-boosting (titer 44) and a 7-fold increase over the levels of both Day 57 (titer 104) and the human convalescent sera, HCS (titer 102).

**Figure 4.**
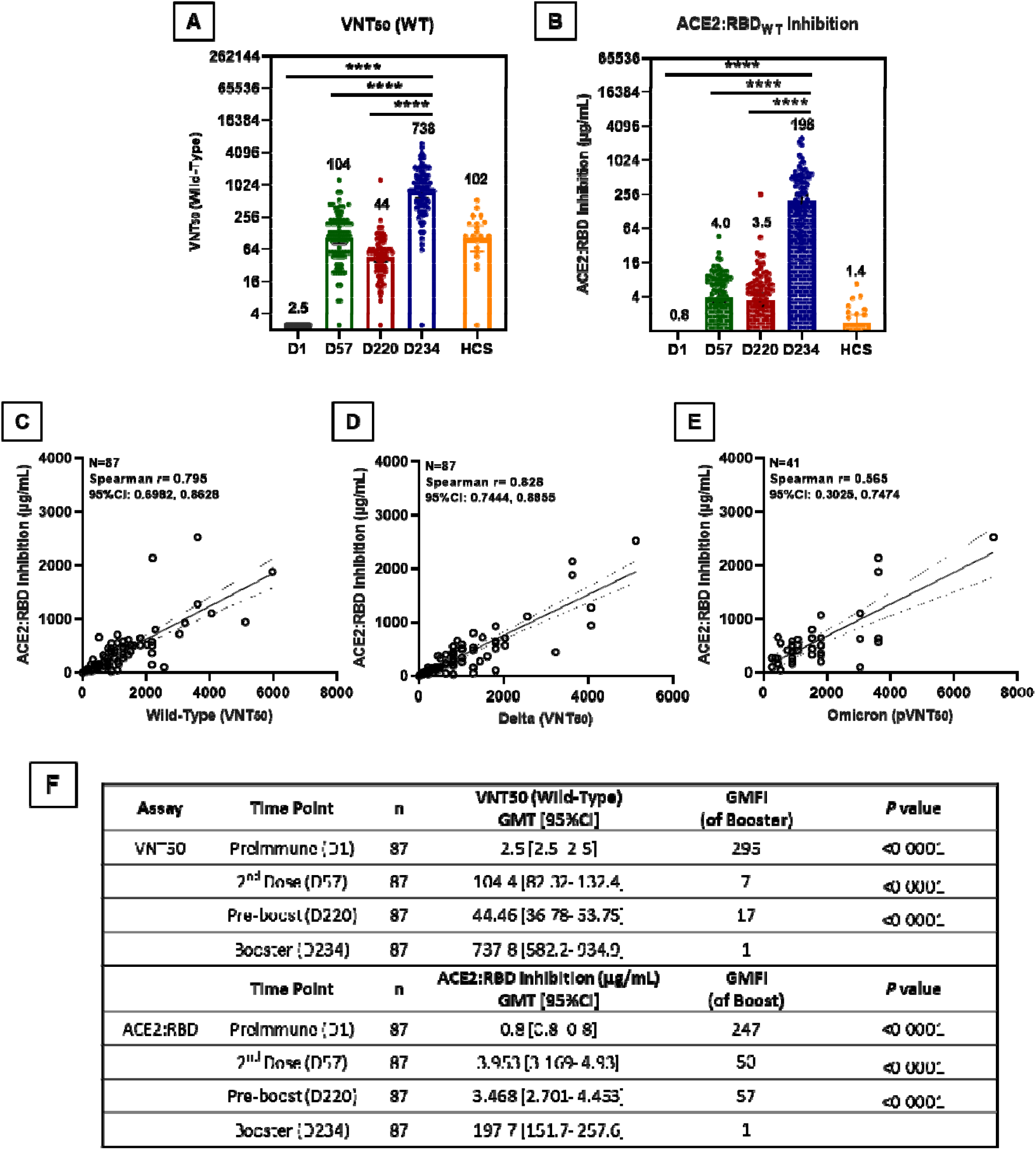
Functional correlations between ACE2:RBD_WT_ binding inhibition and viral-neutralization. Of 871 participants enrolled in the Phase-2 primary 2-dose series and grouped for Immunogenicity investigation, serum samples from 87 participants who had received a booster 3^rd^-dose of 100 μg UB-612 were collected at Day 1 (pre-dose 1), Day 57 (28 days post-dose 2), Day 220 (pre-booster between Days 197 to 242,), Day 234 (14 days post-booster between Days 211 to Day 256). HCS from 20 SARS-CoV-2 infected individuals were also included for comparative testing by two functional assays: **(A)** viral-neutralizing titer (VNT_50_) against live wild-type Wuhan strain (WT) by CPE method; and **(B)** the antibody concentration calibrated with an internal standard for ACE2:RBD_WT_ binding inhibition by ELISA. The correlations are explored between the two function assays, i.e., ACE2:RBD_WT_ binding inhibition ELISA and the viral-neutralizing titers against the live virus (VNT_50_ for original wild-type and Delta strains) or the psuedovirus (pVNT_50_ for Omicron strain). The RBD_WT_ stands for the RBD binding protein domain bearing amino acid sequence of the original SARS-CoV-2 wild-type (WT) Wuhan strain. The correlations were explored for **(C)** ACE2:RBD_WT_ inhibition vs. anti-WT VNT_50_, **(D)** ACE2:RBD_WT_ inhibition vs. anti-Delta VNT_50_, and **(E)** ACE2:RBD_WT_ inhibition vs. anti-Omicron pVNT_50_. The correlation coefficients were evaluated by Spearman r with 95% CI. Statistical analysis was performed with the Student’s t-test (ns, *p*>0.05; ***, *p*<0.001; ****, *p*<0.0001). **(F)** Summary of geometric mean titer (GMT) and 95% CI are presented for plots as shown in Figures (**A**) and (**B**).

A striking post-booster functional antibody-mediated inhibition of ACE2:RBD_**WT**_ binding on ELISA was notable at a high titer (expressed in standard-calibrated antibody concentration) of 198 μg/mL (**Figure 4B**), a ∼57-fold increase over both the pre-boosting titer of 3.5 μg/mL and the Day 57 titer of 3.5 μg/mL, and a profound 140-fold over the HCS titer of 1.4 μg/mL. As the ELISA methodology measures neutralizing (inhibitory) antibodies against RBD_**WT**_ binding to ACE2 receptor, the low HCS titer suggests that the majority of antibodies in HCS appears to bind more to the allosteric sites (N- or C-terminal domain of the S1) than to the orthosteric (RBD) sites.

Of note, UB-612 induces a durable neutralizing antibody titer level, observed between Day 57 (post-2^nd^ dose) vs. Day 220 (pre-boosting), a 42% retainment for VNT_50_ (titer, 104 vs. 44) against live WT virus (**Figure 4A**) and a 88% retainment for ACE2:RBD_**WT**_ (μg/mL, 4.0 vs. 3.5) binding inhibition (**Figure 4B**).

The neutralization of ACE2:RBD_**WT**_ binding on ELISA correlates well with both anti-WT (**Figure 4C**) and anti-Delta VNT_50_ (**Figure 4D**) findings, both showed a similar high correlative Spearman r = 0.795 and 0.828, respectively. A lesser but significant correlation exists for ACE2:RBD_WT_ binding inhibition and anti-Omicron pVNT_50_, with a Spearman r = 0.565 (**Figure 4E**).

## DISCUSSION

The present Phase-2 extension study (participants aged 18-85 years) has shown that UB-612 booster (the third dose) is safe and well tolerated (**eFigure 3 in the Supplement**), and can induce unusually high levels of cross-reactive neutralizing titers against the Delta and Omicron variants (**Figure 3; Table 1**) that parallel with robust, cross-reactive VoC antigen-specific T cell immunity (**Figure 2**). In particular, the booster can induce essentially the same striking degree of neutralizing strength against Delta or Omicron for both the elderly and the young adults (**Figures 3B and 3D**); and the high viral-neutralizing titers bear only a modest ∼1.0-fold (anti-Delta) to 5.0-fold (anti-Omicron) GMFR relative to WT virus (**Figure 3**). Similar fold-reduction landscape is obvious in the primary and booster series in the Phase-1 trial (participants aged 20-55 years),^24^ across all VoCs and other Variants of Interest (**eFigures 5 and 6 in the Supplement**).

**Table 1.** Viral-neutralizing antibody titers against SARS-CoV-2 wild-type (WT) and Omicron variant upon homologous boosting.

| Vaccines<sup>a</sup><br>Homo-booster | Booster time<br>(post-2<sup>nd</sup> dose) | NeuAb assay<br>(Method/Unit) | WT<br>(GMT)<sup>b</sup> | Omicron<br>(GMT)<sup>c</sup> | WT/Omicron<br>(GMFR)<sup>d</sup> |
| --- | --- | --- | --- | --- | --- |
| UB-612 | 6-8 months | PNA/pVNT<sub>50</sub> | 6245 | 1196 | 5.2 |
| mRNA-1273 | 9 months | PNA/ID<sub>50</sub> | 4216 | 650 | 6.5 |
| BNT162b2 | >6 months | PRNT/PRNT<sub>50</sub> | 368 | 164 | 2.2 |
| MVC-COV1901 | 6 months | PNA/ID<sub>50</sub> | 1280-640 | 160-80 | 8.7 |
| AZD1222 | 6 months | FRNT/FRNT<sub>50</sub> | 726 | 57 | 12.7 |
| BBIBP-CorV | 8-9 months | PNA /pVNT<sub>50</sub> | 295 | 15 | 20.1 |

On safety, no concerns of vaccine-related SAE were identified (with ∼4000 individuals vaccinated so far). While UB-612 has not yet been deployed geographically wide enough, its peptide-protein subunit composition containing aluminum and CpG as the adjuvants (**Figure 1A**) suggests that it should have a high safety profile, unlike mRNA and adeno-vectored vaccines that, upon repeat dosing, could have more severe adverse events including rare but serious adverse reactions such as myocarditis, pericarditis, Guillain-Barre syndrome, and thrombosis-thrombocytopenia.^25^

On cellular immunity, UB-612 can induce robust and long-lasting Th1 cell responses and prompt a high level of memory recall upon boosting. In the Phase-1 primary series, the post-2^nd^ dose PBMCs stimulated RBD/Th/CTL can induce high antigen-specific Th1-dominant IFN-γ^+^− ELISpot T cell responses at 254 SFU/10^6^ cells and a long-lasting response level with ∼50% retained (the pre-boosting 121 vs. peak 254) >6 months post-2^nd^ dose.^24^ In the Phase-2 primary series, a higher SFU units of 374 post-2^nd^ dose and of 261 >6 months later (pre-boosting) (**Figures 2A and 2D**) represents even a higher 70% (261 vs. 374) retainment of T cell immunity, with the SFU surged to 444 post-booster. The minor Th2 IL-4^+^-T cell response found in the Phase-2 trial (**Figures 2B and 2E**) reconfirms that observed in the Phase-1 study.^24^

Evidently, the Th/CTL peptide pool (containing N, M, and S2 epitope peptides) is the principal driver of T cell immunity, with SFU units at 322 post-2^nd^ dose and maintained at 317 post-booster (**Figures 2A and 2D**). Furthermore, a strong, durable cytotoxic (CD107a^+^-Granzyme B^+^) activity of CD8^+^ T lymphocytes (CTL) accounting for a high CTL frequency at 3.6% post-2^nd^ dose and persists at 1.8% pre-booster (**Figures 2C and 2F**) indicates a 50% retainment of CTL activity more than 6 months post-2^nd^ dose, which maintains at the same level observed 14 days post-booster.

The magnitude of the UB-612 booster-recalled T cell immunity in the present Phase-2 homologous-boosting (SFU units range ∼320-450) was found to be greater than any of those produced under “mix-and-match” heterologous-boosting settings by the currently authorized mRNA, adeno-vectored, and Spike protein-based vaccines, which reported SFU/10^6^ PBMC units (stimulated with spike peptide-specific to WT, Delta and Beta) to be in the range of 40 to 150.^26^ That the rationally-designed UB-612 S1-RBD-sFc protein-subunit vaccine is armed with T cell immunity-promoting N (nucleocapsid), M (membrane) and S2 (spike) epitope peptides (**Figure 1B**), which are highly conserved across all VoCs, may underlie the UB-612’s differentially pronounced booster-induced T cell immunity.

The much more booster-enhanced T-cell immunity via UB-612 is also supported by the development of a plain T-cell vaccine containing a six-peptide backbone that, as a T-cell booster, triggered dramatic multifunctional CD4 and CD8 T-cell responses,^27^ which showed notable benefits to B-cell deficient, immunocompromised patients who could not mount B-cell antibody responses. This raises the concerns over the fact that humoral antibody response has long been used as a sole metric of protective immunity,^28,29^ which lacks full understanding of human post-vaccination immune responses as antibody response is generally shorter-lived than virus-reactive T cells.^30-32^

Further, the SARS-CoV-2’s non-spike structure proteins of envelope (E), membrane (M) and nucleocapsid (N) are the regions critically involved in the host cell interferon response and T-cell memory.^19-22^ These structural proteins of virus’ main body when presented would fall beyond recognition by the currently authorized vaccines that based on the outer spike only, implicating that these vaccines bear an intrinsic shortfall,^33^ lacking at least N- and M-specific T cell immunity. The adaptive immune response is a major determinant of the clinical outcome, to which T cell immunity plays a central role in the control of SARS-CoV-2 infection and its importance have been underestimated thus far.^34^ In fact, the non-spike viral structure proteins have long been grossly overlooked since the development of COVID vaccines.

On humoral immunity, UB-612 booster shots in both Phase-1 and Phase-2 studies exhibit similar high-titer profiles. The Phase-1 post-2^nd^ dose booster (adults aged 20-55 years) induced unusually high cross-neutralizing anti-Delta VNT_50_ titers against live viruses (WT 3,992 vs. Delta at 2,358; 1.7-fold reduction) (**eFigure 6A in the Supplement**),^24^ and the Phase-2 booster vaccination (adults aged 18-85 years) elicited pronounced VNT_50_ titers as well (WT at 1,711 vs. Delta at 1,282; 1.3-fold reduction) (**Figure 3C**). In addition, although not investigated in the Phase-2 study, we have found in the phase-1 primary series that UB-612 could induce a long-lasting viral-neutralizing antibody against WT strain with a half-life of 187 days.^24^

Potent post-booster anti-Omicron effects (pVNT_50_) against pseudovirus are notably similar: WT at 12,778 vs. Omicron at 2,325 with 5.4-fold reduction in Phase-1 (**eFigure 6B in the Supplement**)^24^; and WT 6,245 vs. 1196 with 5.2-fold reduction in Phase-2 (**Figure 3A**). Though with a ∼5-fold reduction for Omicron in both clinical studies, the neutralizing pVNT_50_ in the range of 1,196 to 2,325 are of high potency. Comparatively, the differential fold reductions reveal that UB-612 may neutralize Delta with a potency around 3- to 4-fold greater than against Omicron.

Furthermore, there have been reports on homologous booster vaccination by other vaccine platforms (**Table 1**),^35-39^ where the post-booster anti-WT pVNT_50_ were reported to range from the low 295 to the high 6,245 (UB-612), and the anti-Omicron to range from the low 15 to the high 1,196 (UB-612). The UB-612’s anti-Omicron pVNT_50_ profile in contrast to other vaccine platforms is in good agreement with the finding of high performance of the UB-612 booster combating live WT and live Delta virus strains observed in the Phase-1 booster vaccination.^24^ In contrast to other vaccine platforms, anti-WT VNT_50_ titers were reported to range from the low 122 to the high 3,992 (UB-612), and the anti-Delta to range from the low 54 to the high 2,358 (UB-612). UB-612 appears to bear a competitive edge over other vaccine platforms in neutralizing both Omicron and Delta variants.

We also observed significant pVNT_50_ titers against pseudovirus of SARS-CoV-2 variants for the Phase-1 post-2^nd^ dose serum (28 days post-2^nd^ dose), which ranges from 76.6 for Beta, 224 for Gamma, 246 for Delta, 374 for Alpha, and 394 for WT (**eFigure 5 in the Supplement**), representing an 1.0 to 5.1-fold reduction relative to the WT, suggestive that UB-612 could be effective against all VoCs.

The profound post-booster neutralization effect against both live WT and live Delta, and pseudovirus WT and Omicron variants, illustrates one unique feature of UB-612, i.e., the immune response is directed solely at the binding domain (RBD) that reacts with ACE2 receptor. The RBD-focused design leaves little non-conserved sites on the spike for viral mutation, thus boosting promptly recalls of high levels of both viral-neutralizing (**Figure 3**) and RBD-ACE2 binding inhibition antibodies (**Figure 4A**), and both functional activities are significantly inter-correlated (**Figure 4B)**. Of note, in this Phase-2 primary series, the anti-WT VNT_50_ (**Figure 4A**) and the ACE2:RBD binding inhibition titer (**Figure 4B**) are durable over the Day 57 and Day 220.

The finding that the UB-612 induced much higher fold-increases (GMRIs) in blocking the RBD-ACE2 interaction than that by HCS (**Figure 4B**) suggests that most of the antibodies in HCS may bind allosterically to the viral spike (N- or C-terminal domain of the S), rather than orthosterically to the RBD sites and may include some unwanted enhancing antibodies. This warrants further investigation including sera from re-infections and breakthrough infections from all vaccine platforms.

Booster vaccination can reduce rates of hospitalization and severe diseases, yet offers less protection against infection and mild disease.^1-11^ The pathologically-lesser but hyper-transmissible Omicron shall not be treated as a trivia as the coming of flu. COVID by itself can take a serious long-term toll on heart health^12^, presumably to stay as a part of long-haul COVID.^13^ This toll, beyond the already known myocarditis and pericarditis associated with mRNA vaccines,^14^ encompasses a cluster of inflammatory cardiovascular disorders that elevates, depending on COVID severity, from asymptomatic, symptomatic, to acute infection cases.^12,15,16^ Facing ever-emergent variants and long-haul COVID, urgent development of composition-updated new vaccines that can enhance immunity with sufficient magnitude, durability and breadth of virus coverage has been strongly advocated.^17,18^

As memory B and T cells are critical in secondary responses to infection, a successful vaccine must generate and maintain immunological memory,^40,41^ and to mount a rapid recall of effective humoral and cellular responses upon natural exposure or vaccine boosting. By rational design, UB-612 vaccine product has demonstrated such important vaccine features of balanced B- and T-immunity through these clinical studies.

In particular, the UB-612-induced T cell immunity would comprehensively recognize spike (S2) and non-spike structure N (nucleocapsid) and M (membrane) proteins, which may boost the potential in favor of viral clearance of the infected cells regardless of mutations found in Delta and Omicron, as their mutation sites are not to overlap any of the amino acid residues on the precision-designed S2, N, and M epitope peptides that are highly conserved (or rarely mutate) across all VoCs^23^ (**Figure 1B**). Presumably, the viral proteins on Omicron that are substantially conserved can serve as strong T-cell activators and induce long-lasting T-cell response^42^, which can synergize with B-cell memory for enhanced immunity. And, as non-spike structure N and M proteins fall beyond recognition by the currently authorized COVID vaccines, UB-612 may serve as a universal vaccine to fend off new Variants of Concern such as Delta, Omicron and other ever-emergent SARS-CoV-2 variants with durable and balanced B-and T-cell immunity.

### Limitation

This study has four limitations. First, UB-612 has not yet been widely deployed geographically. Second, in-depth biomarker analysis beyond those described in this report was not conducted due to insufficient volume of blood and samples retained. Third, we used Th/CTL peptide pool for an overall in vitro assessment of immune enhancement upon booster without delineation as to which peptide contributes most to mounting T-cell immunity in each of the vaccinees, though pooling may produce synergistic effect. Fourth, the post-booster functional analyses were carried out for a short period of time, which lacks information as to how durable the B- and T-cell immune response would last. However, it is worthy to note that UB-612 multitope subunit vaccine is the first designer vaccine to elicit a potent balanced B- and T-cell immunity against SARS-CoV-2 infection. Development of a new generation of such B-T combined vaccines has been explored. Large scale efficacy trials in geographically diverse areas enrolling young and elderly with various comorbidities, in both homologous and heterologous booster studies, are undergoing to accelerate the vaccine development for its timely introduction to prevent and control COVID.

## CONCLUSIONS

The UB-612-induced T cells would comprehensively recognize spike (S2) and non-spike structure N (nucleocapsid) and M (membrane) proteins that may seed the potential favorable for viral clearance once infection occurs, regardless of mutations in the spike or non-spike protein domains on Delta and Omicron (**Figure 1B**). As non-spike structure N and M proteins fall beyond recognition by the currently authorized COVID vaccines, UB-612 may serve as a universal vaccine to ward off VoCs including Delta, Omicron and ever-emergent SARS-CoV-2 variants with a durable and balanced B-and T-cell immunity.

## Data Availability

All data produced in the present work are contained in the manuscript

https://unitedbiomedical.com

## AUTHOR CONTRIBUTIONS

CYW, KPH, HKK, HL, YHS and WJP, were responsible for vaccine development including vaccine formulation design, protocol design and implementation of the clinical studies, acquisition and interpretation of the clinical data. HKK, KLH, JC, MSW, YTY, were responsible for assay development and validation, laboratory testing and data collection, and preparation of respective reports. CYW, YHS, HKK, HL, BSK and WJP had full access to and verified all the data in the study and take responsibility for the integrity and accuracy of the data analysis. BSK and CYW drafted and prepared the manuscript. All authors reviewed and approved the final version of the manuscript. CYW had final responsibility for the decision to submit for publication.

## DATA SHARING

The study protocols are provided in the Supplemental Appendices. Individual participant data will be made available when the trial is complete with data to be shared through a secure online platform.

## ACKNOWLEDGEMENTS

The study was funded by UBI Asia (study sponsor) and the Taiwan Centers for Disease Control, Ministry of Health and Welfare. The sponsor co-designed the trial and coordinated interactions with contract Clinical Research Organization (CRO) StatPlus staff and regulatory authorities. The CRO took charge of trial operation to meet the required standards of the International Council for Harmonization of Technical Requirements for Pharmaceuticals for Human Use and Good Clinical Practice guidelines. The Independent Data Monitoring Committee (IDMC) oversaw the safety data and gave recommendations to the sponsor. The interim analysis was done by the CRO StatPlus. We thank all the trial participants for their dedication to these trials; the investigation staff at China Medical University Hospital, Taipei Medical University Hospital, Far Eastern Memorial Hospital, National Cheng Kung University Hospital, Linkou Chang Gung Memorial Hospital, Kaohsiung Chang Gung Memorial Hospital, Kaohsiung Medical University Hospital, Tri-Service General Hospital, Taipei Veterans General Hospital, Kaohsiung Veterans General Hospital, Changhua Christian Hospital and Taichung Veterans General Hospital for their involvement in conducting the trial; and members of the IDMC for their dedication and guidance.

Special thanks are also extended to the clinical associates from StatPlus, Inc and UBI Asia; the CMC task forces from both United BioPharma, Inc. and UBI Pharma, Inc; and team members at Institute of Biomedical Sciences, Academia Sinica for the live virus neutralization assay; and team members at the RNAi Core Facility, Academia Sinica for the pseudovirus neutralization assay. All health convalescent sera were supplied by Biobank at the National Health Research Institutes (NHRI), Taiwan.

Finally, critical review of the manuscript by professor Chwan-Chuen King of Institute of Epidemiology and Preventive Medicine, College of Public Health, National Taiwan University, and special administrative support by Jalon Tai, Liang Kai Huang, Peter Hu and Fran Volz from the UBI group are acknowledged with gratitude.

