## Supplementary Materials for "UB-612, a Multitope Universal Vaccine Eliciting a Balanced B and T Cell Immunity against SARS-CoV-2 Variants of Concern"

Chang Yi Wang, on assignment at UBI Asia, Hsinchu, Taiwan

**Key words:** UB-612, Multitope Universal Vaccine, booster vaccination, SARS-CoV-2, Sarbecovirus, and Variants of Concern.

**Supplemental Information**

**Supplementary Figures**

Figure S3. Incidence of adverse effects in the Phase-2 primary 2-dose and extended booster third-dose series……………………………………………………………………….5

Figure S4. Immunogenicity overview (pre- and post-booster) on homologous boosting………...6

Figure S6. Viral-neutralizing titers against live SARS-CoV-2 wild type (Wuhan) and Delta

variant (VNT_50_), and pseudo SARS-CoV-2 wild type (Wuhan) and Omicron

variant (pVNT_50_) after the booster third-dose in the Phase-1 trial…………………….8

**Supplementary Appendices** (available upon request)

Appendix 1

Phase 2 study V-205 protocol

Appendix 2

Phase 2 study V-205 IRB approval letter 1

Phase 2 study V-205 IRB approval letter 2

Phase 2 study V-205 IRB approval letter 3

Appendix 3

Phase 2 study V-205 Informed Consent Form (ICF)

Appendix 4

CONSORT checklist

**SUPPLEMENTARY FIGURES**

**Figure S****1. Distribution of SAES-CoV-2 Delta variant is being replaced**

**by the Omicron variant**

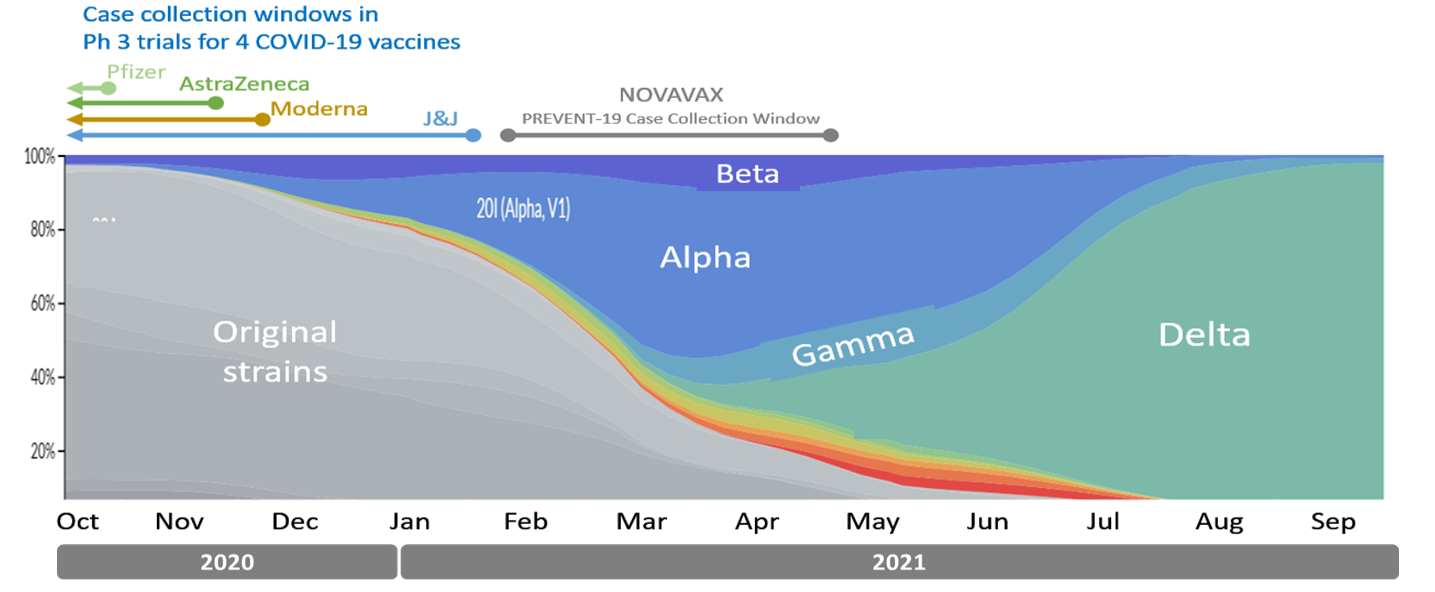

**A**

*Source: Adapted from Strain Sistribution data from Nextstrain.org (GISAID data) as of 04-Oct-2021,*

*with edits for vaccines EUA-authorised and (Variants of Concern).*

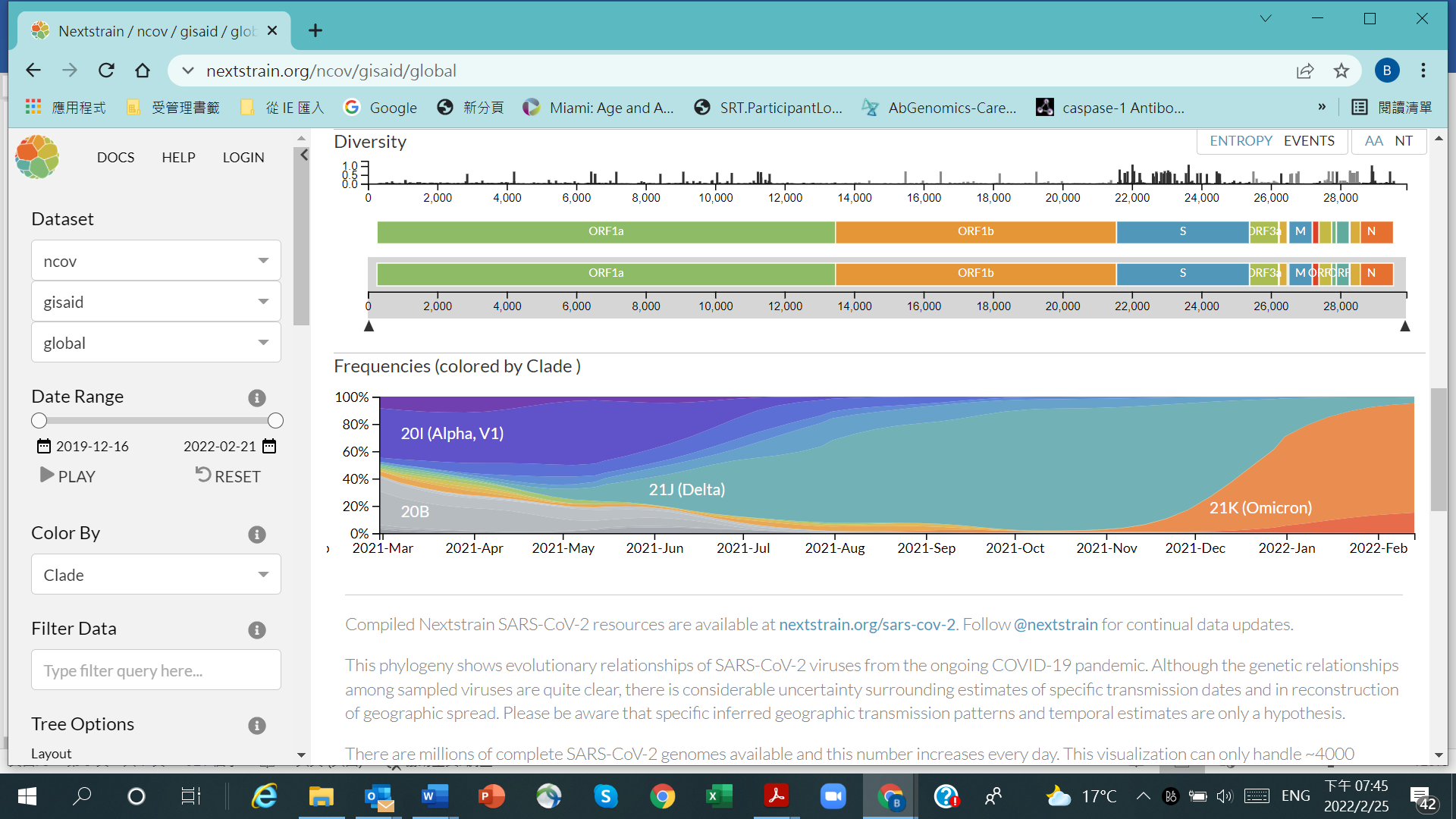

**B**

*Source: Adapted from Strain Distribution data from Nextstrain.org (GISAID data) as of Feb-13-2022.*

The changes overtime in % distribution of SARS-CoV-2 Variants of Concern (VoCs). **(A)** The time points when the five COVID-19 vaccines (Pfizer, Moderna, AZ, J&J and Novavax) were developed and EUA authorized; and the Delta variant that dominated with >95% of all infection cases observed in October 2021. **(B)** The once globally-dominant Delta variant in infectivity is being overtaken and replaced by the heavily-mutated Omicron strain, which constitutes 95% of all infection cases worldwide (80% 21K, BA.1; 15% 21L, BA.2), as of Feb. 13, 2022.

**Figure S****2. Flow** **of UB-612 phase-2 primary 2-dose series with extension booster**

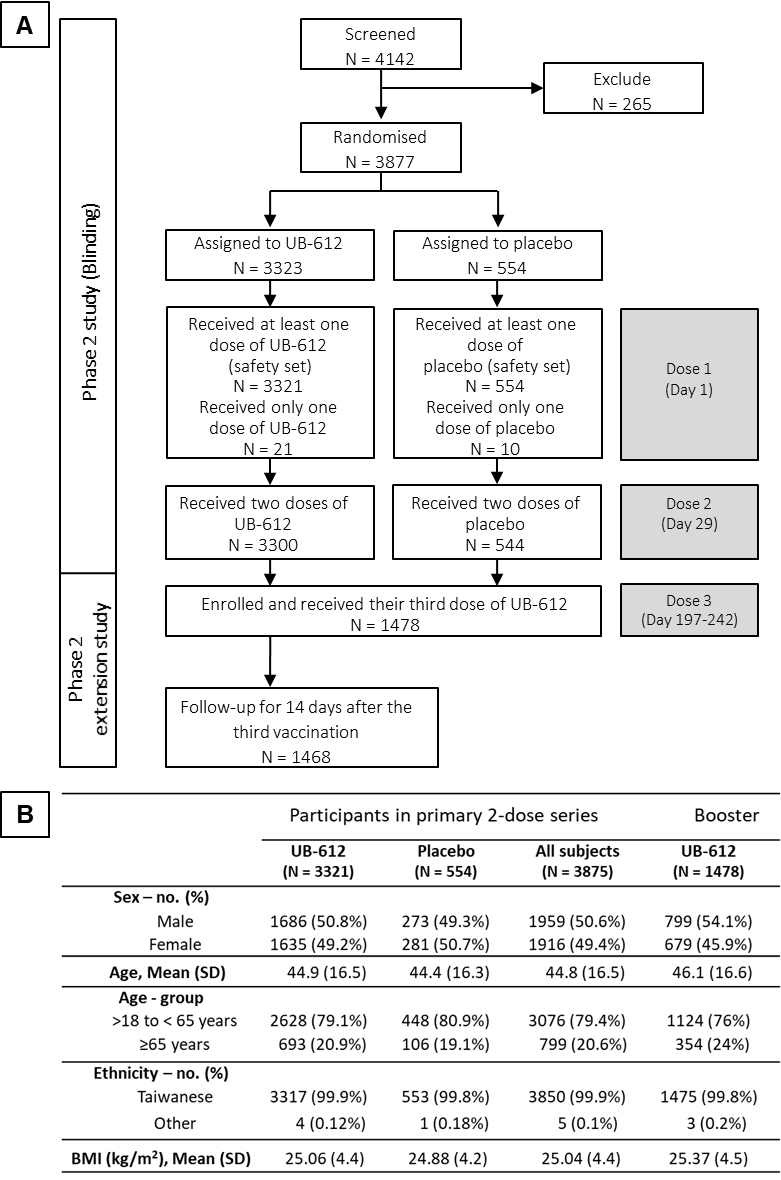

The study design of the Phase-2 primary 2-dose series (100 μg dose; 28 days apart) of UB-612; and the extension study of booster vaccination [NCT04773067] conducted between Oct. 16, 2021 and Apr. 16, 2022. **(A)** Of the primary series (n = 3875), a total of 1,478 participants (aged at 18-85 years) were enrolled to receive the booster third-dose of 100 μg UB-612; **(B)** the characteristics of the study participants in the primary and booster series.

**Figure** **S3. Incidence of adverse effects in the Phase-2 primary 2-dose and extended booster third-dose series**

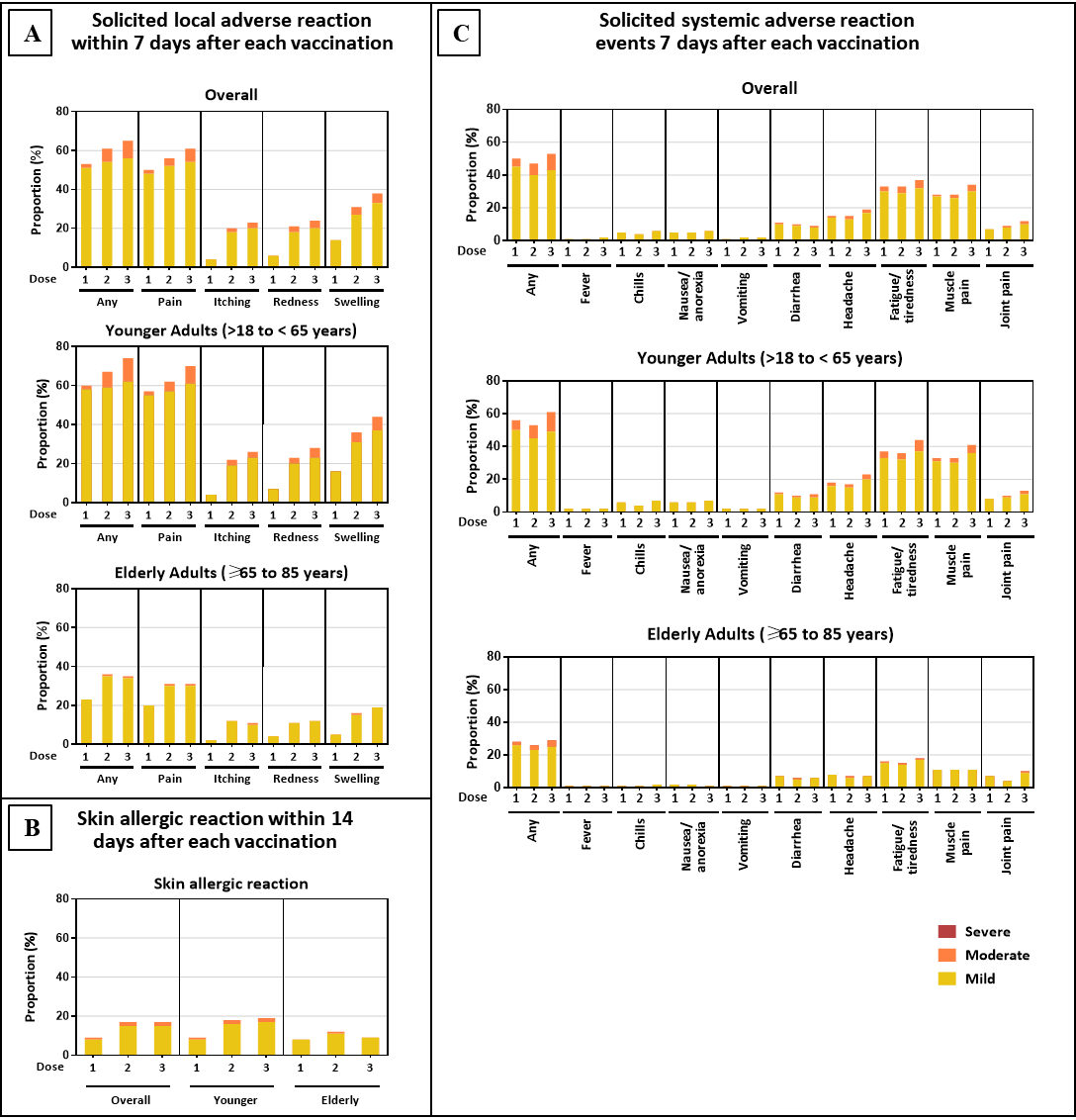

**(A)** Solicited local adverse reaction within 7 days after each vaccination. **(B)** Skin allergic reaction within 14 days after each vaccination. **(C)** Solicited systemic adverse reaction events 7 days after each vaccination (Doses 1 and 2 in the primary series; Dose 3 as a booster)

**Figure S4. Immunogenicity overview (pre- and post-booster) on homologous boosting**

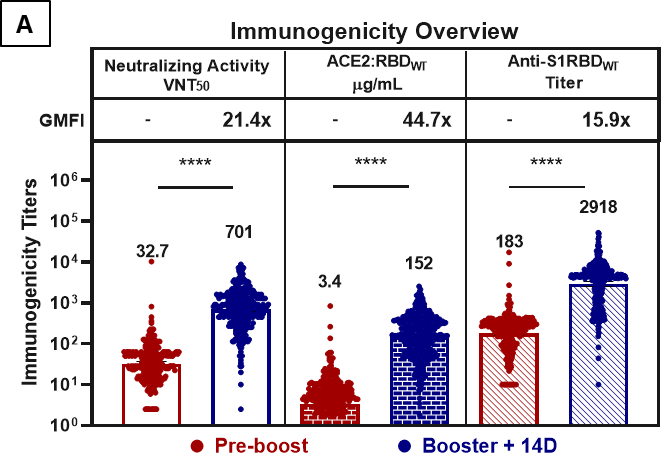

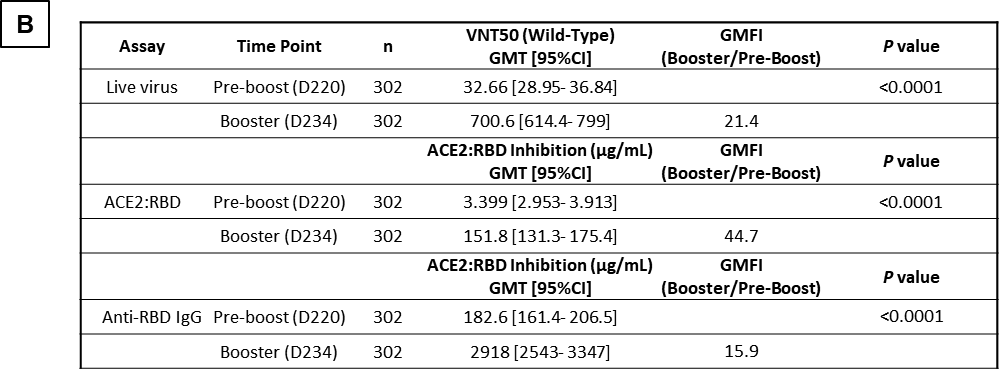

Immunogenicity overview of 302 participants (n = 208 for aged 18-65 years; n = 94 for aged 65-85 years) received a booster 3^rd^-dose. The serum samples of 302 participants were collected at the indicted time points, Days 197 to 242 (the pre-booster day) and Days 211 to 256 (14 days post-booster), and tested for neutralizing antibody levels that inhibit 50% of live SARS-CoV-2 wild-type (WT, Wuhan strain), the inhibitory titers against S1-RBD binding to ACE2 by ELISA, and anti-S1-RBD IgG antibody titers by ELISA. The data were expressed as geometric mean titers GMT and 95% CI. Statistical analysis was performed by the Student’s t-test (ns *p*>0.05, **** *p*<0.0001). **(B)** Summary of geometric mean titer (GMT) with 95% CI are presented for plots shown in Figure **(A)**.

**Figure S5.** **Neutralization of different SARS-CoV-2 variants by sera (Day 42 and Day 56)**

**from the Phase-1 primary series**

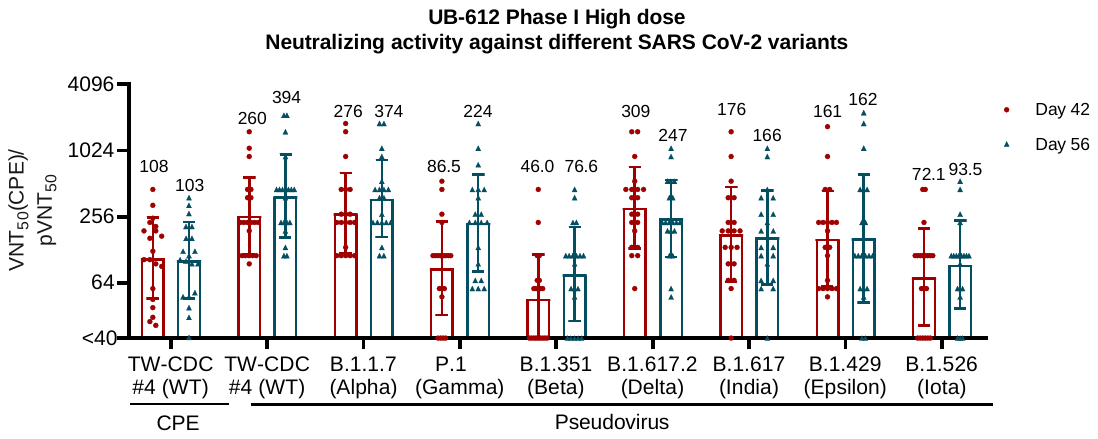

**A**

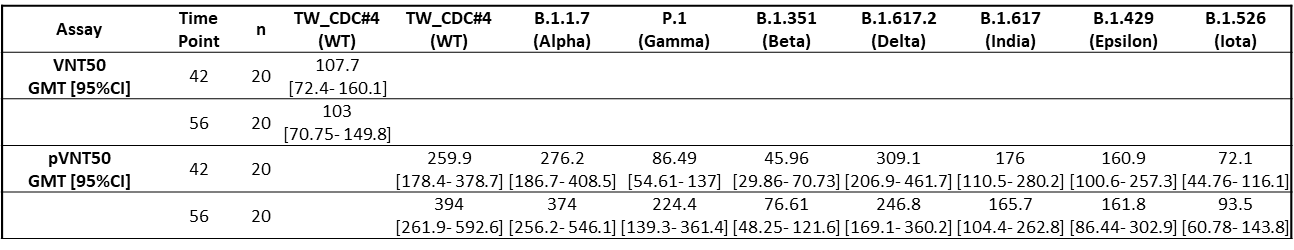

**B**

Ability of sera of UB-612 vaccinated subjects to neutralize SARS-CoV-2 variants in a WT CPE assay or a pseudovirus assay. Neutralization titers (Day 42 and Day 56 after UB-612 vaccination) of seropositive vaccinees (n = 20) from Phase-1 high dose 100-μg cohort are tested for TW-CDC#4 (WT), variants B.1.1.7 (Alpha), P.1 (Gamma), B.1.429 (Epsilon), B.1.526 (Iota), B.1.351 (Beta) B.1.617 (India), and B1.617.2 (Delta) variants are presented. Values are indicated as geometric mean titres (GMTs). **(A)**Neutralization titre <40 was plotted as 20 by the viral-neutralizing titers (pVNT_50_) based on a pseudovirus assay. The reduction fold for each virus variant relative to wild-type WT (TW-CDC #4) were calculated to be approximately 1.1 for B.1.1.7 (Alpha), 1.8 for P.1 (Gamma), 5.1 for B.1.351 (Beta), 1.6 for B.1.1617.2 (Delta), 2.4 for B.1.617 (India), 2.4 for B.1.429 (Epsilon), and 4.2 for B.1.526 (Iota). The reduction fold for the variant strains including VoCs are in the range of 1.0 to 5.0. **(B)** Summary of geometric mean titers (GMT) are presented with 95% CI for plot shown in Figure (**A)**.

**Figure S6. Viral-neutralizing titers against live SARS-CoV-2 wild type (Wuhan) and Delta**

**variant (VNT_50_), and pseudo SARS-CoV-2 wild type (Wuhan) and Omicron**

**variant (pVNT_50_) after the booster third-dose in the Phase-1 trial**

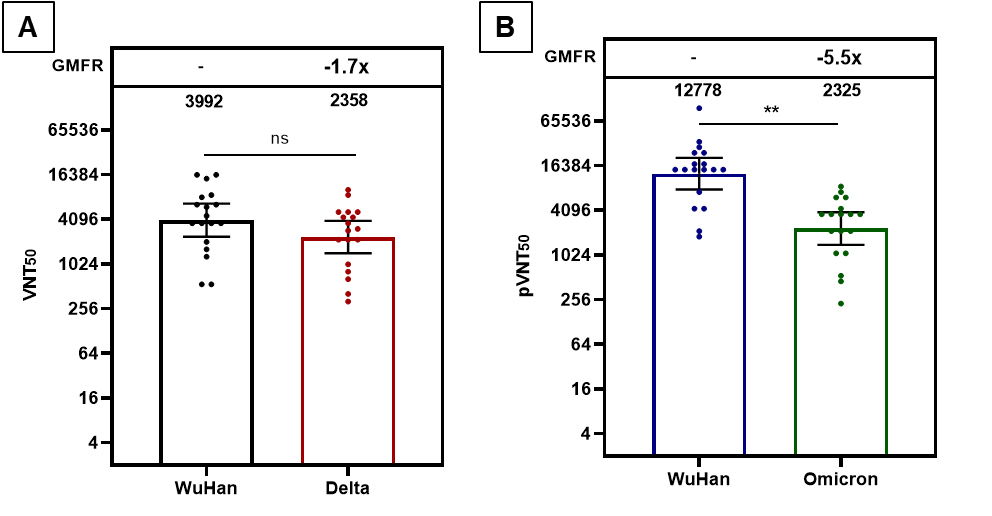

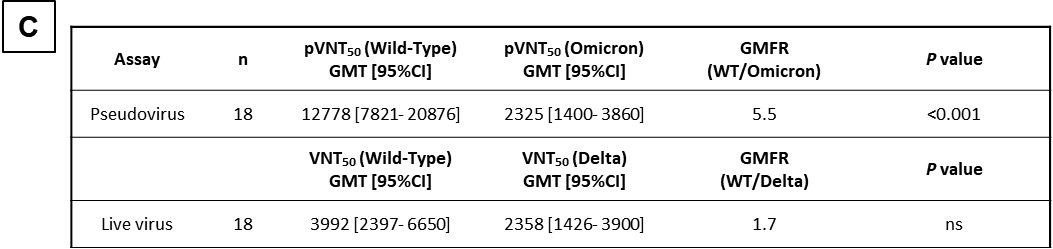

Geometric mean titers (GMT) at 50% viral-neutralization observed 14 days after the booster third-dose of 100-µg administered at mean Day 286 (Days 255-316) after the primary 2-dose series (Days 0 and 28) of the 196-day Phase-1 trial. **(A)** In the participants of the 100-µg group (n = 18) with healthy adults aged at 20-55 years, the post-booster VNT_50_ titer reached at 3,992 against live SARS-CoV-2 Wuhan wild-type, and at 2,358 against live Delta variant. **(B)** Similarly, unusually high post-booster pVNT_50_ against Wuhan wild-type pseudovirus at 12,778, and at 2,325 against Omicron variant. **(C)** Summary of geometric mean titer (GMT) with 95% CI are presented for plots shown in Figures **(A)** and **(B)**.
